# SPECIFIC DETECTION OF SARS-COV-2 B.1.1.529 (OMICRON) VARIANT BY FOUR RT-qPCR DIFFERENTIAL ASSAYS

**DOI:** 10.1101/2021.12.07.21267293

**Authors:** Oran Erster, Adi Beth-Din, Hadar Asraf, Virginia Levy, Areej Kabat, Batya Mannasse, Roberto Azar, Ohad Shifman, Shirley Lazar, Michal Mandelboim, Shay Fleishon, Ella Mendelson, Neta S Zuckerman

**Affiliations:** Central Virology Laboratory, Public Health Services, Ministry of Health, Chaim Sheba Medical Center, Ramat Gan, Israel; Israel Institute for Biological Research; School of Public Health, Sackler Faculty of Medicine, Tel-Aviv University, Tel-Aviv, Israel

**Keywords:** SARS-COV-2, B.1.1.529 VOC Omicron, RT-qPCR, differential assay

## Abstract

In this report, we describe four RT-qPCR assays that enable rapid identification of the newly emerging SARS-COV-2 Omicron (B.1.1.529) variant of concern. The assays target Omicron characteristic mutations in the nsp6 (Orf1a), spike and nucleocapsid genes. We demonstrate that the assays are straightforward to assemble and perform, are amendable for multiplexing, and may be used as a reliable first-line tool to identify B.1.1.529 suspected samples. Importantly, this is a preliminary development report. Further validation and optimization of the assays described herein will be published hereafter.

## 1. Introduction

The SARS-COV-2 (SC-2) B.1.1.529 (Omicron) variant recently emerged in South Africa and Botswana in November 2021, and was identified in numerous countries worldwide shortly after, with travel-related cases identified in Hong Kong, Belgium and Israel at the end of November. The Omicron variant includes 30 mutations in the spike protein alone, 15 of which are in the receptor binding domain (RBD). Being the most highly divergent SARS-CoV-2 variant yet, this newly emerging variant of concern (VOC) is suspected to have a potential for increased transmissibility and reduced vaccine effectiveness.

Due to the urgent need to provide a first-line approach for specific detection of the Omicron variant, we describe here the preliminary development of four assays that provide rapid identification of suspected Omicron variant samples. The assays were developed by the Israel Ministry of Health Central Virology Laboratory (CVL) and the Israel Institute for Biological Research (IIBR). Some of the reactions in these assays partially rely on established protocols, and enable the detection of mutations that are thus far strongly associated with this variant, based on sequences available in the GISAID database (www.epicov.org/epi3/frontend#5c1e82). The assays were tested with the first Omicron samples identified in Israel and their optimization is currently underway. Since the only circulating variant in Israel is currently B.1.617 (Delta), the specificity of the assays was initially tested using this variant as a reference. In some cases, the assay performance was also tested using the A19 (“WT”) and B.1.1.7 (Alpha) strains. All B.1.1.529 specific reactions were performed in a duplex with the E-sarbeco reaction described by Corman et al. (1), with some modifications that were described previously (2) to render the reaction more suitable for multiplexing. The negative N_del_ reaction is based on the CDC N1 reaction, which is widely used in many diagnostic laboratories (3). The assays described herein can be implemented in any molecular diagnostic laboratory and do not require the use of specific brand instruments, or unique reagents.

## 2. Materials

### 2.1. Primers details

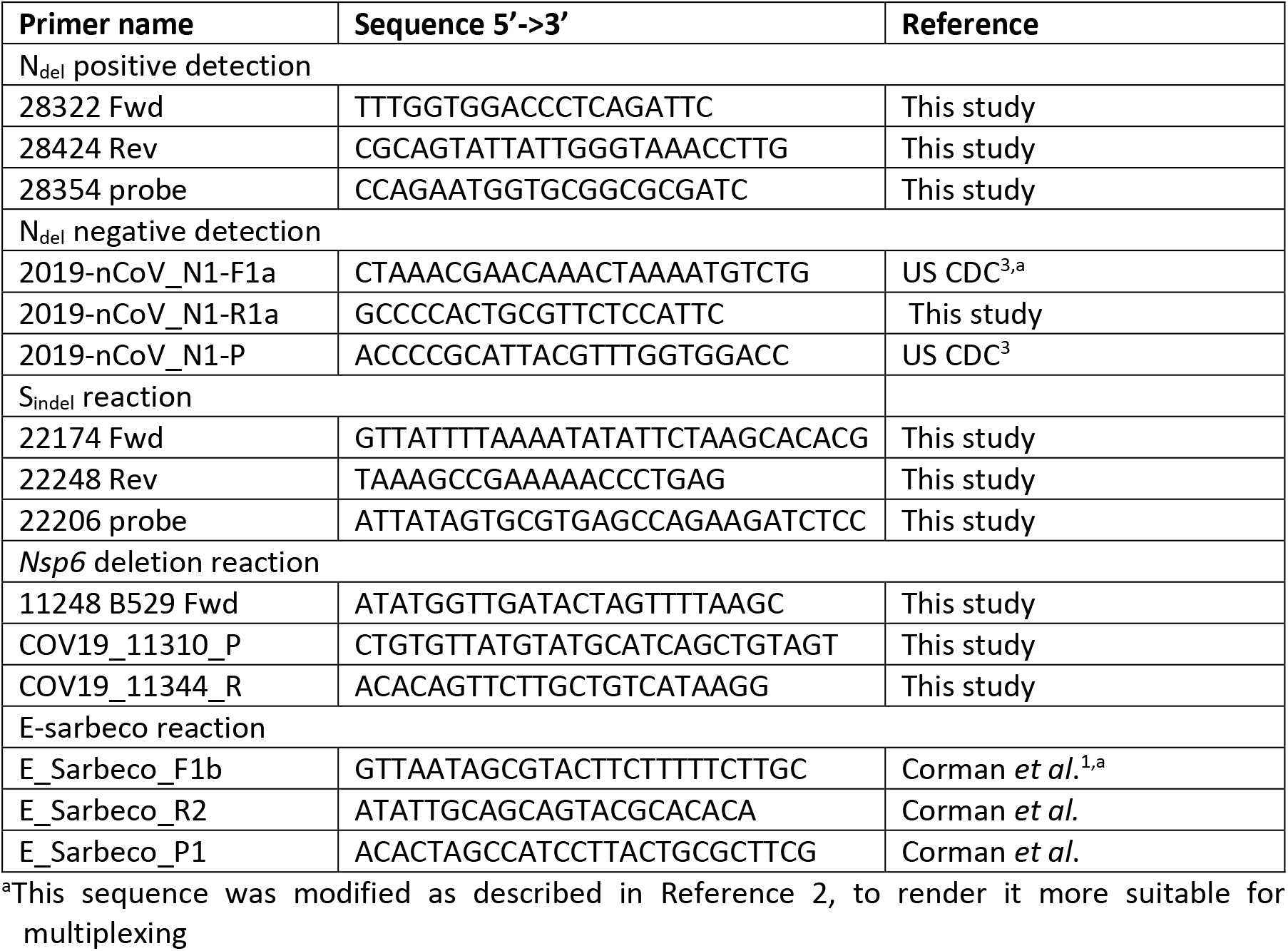

### 2.2. Master mix

The SensiFast Lo-ROX probe 1-step PCR mix was used (*https://www.bioline.com/sensifast-probe-lo-rox-kit.html*).

### 2.3. Positive N_del_ Reaction setup

| Positive $N_{del}$ reaction | | Volume (μl) per reaction |
| --- | --- | --- |
| Master Mix SensiFast Probe Lo-ROX One-Step | Conc / Rxn | 12.5 |
| H<sub>2</sub>O |  | 1.8 |
| E-Sarbeco-F1b 40μM | 600nM | 0.375 |
| E-Sarbeco-R 40μM | 600nM | 0.375 |
| E-Sarbeco-P 20μM FAM | 300nM | 0.375 |
| COV19_28322Fwd_F 40uM stock | 600nM | 0.375 |
| COV19_28412 Rev 40uM | 600nM | 0.375 |
| COV19_22206_P 20uM HEX | 300nM | 0.375 |
| SensiFast RT-Enzyme |  | 0.25 |
| RNAse inhibitor |  | 0.2 |
| Total |  | 17 |
| RNA |  | 8 |

### 2.4. Negative N_del_ reaction setup

2.4. Negative $N_{del}$ reaction setup
| $N_{del}$ negative reaction | | Volume (μl) per reaction |
| --- | --- | --- |
| Master Mix SensiFast Probe Lo-ROX One-Step | Conc / Rxn | 12.5 |
| H<sub>2</sub>O |  | 1.8 |
| E-Sarbeco-F1b 40μM | 600nM | 0.375 |
| E-Sarbeco-R 40μM | 600nM | 0.375 |
| E-Sarbeco-P 20μM FAM | 300nM | 0.375 |
| CDC N1 Fwd 40uM stock | 600nM | 0.375 |
| N1 R1a Rev 40uM | 600nM | 0.375 |
| CDC N1 probe 20uM HEX | 300nM | 0.375 |
| SensiFast RT-Enzyme |  | 0.25 |
| RNAse inhibitor |  | 0.2 |
| Total |  | 17 |
| RNA |  | 8 |

### 2.5. S_22194 indel_ reaction

2.5. $S_{22194}$ *indel* reaction
| $S_{22194}$ *indel* reaction | Volume (μl) per reaction |
| --- | --- |
| Master Mix SensiFast Probe Lo-ROX One-Step | 12.5 |
| H<sub>2</sub>O | 1.8 |
| E-Sarbeco-F1b 40μM | 0.375 |
| E-Sarbeco-R 40μM | 0.375 |
| E-Sarbeco-P 20μM TXRed | 0.375 |
| 22174 Fwd 40μM | 0.375 |
| 22248 Rev 40μM | 0.375 |
| 22206 probe 20μM | 0.375 |
| SensiFast RT-Enzyme | 0.25 |
| RNAse inhibitor | 0.2 |
| Total | 17 |
| RNA | 8 |

### 2.6. NSP6_del_ reaction

| NSP6<sub>del</sub> reaction |  | Volume (μl) per reaction |
| --- | --- | --- |
| Master Mix SensiFast Probe Lo-ROX One-Step | Conc / Rxn | 12.5 |
| H<sub>2</sub>O |  | 1.8 |
| E-Sarbeco-F1b 40μM | 600nM | 0.375 |
| E-Sarbeco-R 40μM | 600nM | 0.375 |
| E-Sarbeco-P 20μM FAM | 300nM | 0.375 |
| 11248 B529 Fwd | 600nM | 0.375 |
| COV19_11344_R | 600nM | 0.375 |
| COV19_11310_P | 300nM | 0.375 |
| SensiFast RT-Enzyme |  | 0.25 |
| RNAse inhibitor |  | 0.2 |
| Total |  | 17 |
| RNA |  | 8 |

### 2.7. Thermal profile

All reactions were run in a Bio-Rad CFX96 instrument, with the same thermal profile, as follows: (1) 45.0°C for 10’10”, (2) 95.0°C for 2’20”, 45X[(3) 95.0°C for 4”, (4) 60.0°C for 20”]. Fluorescence was read at step (4). For all assays except the S_22194 indel_ duplex reaction, fluorescence was read in the FAM and HEX/VIC channels. For the S_22194 indel_ reaction, fluorescence was read in the FAM and Texas-Red (TXRed) channels.

### 2.8 Bioinformatic analysis

Sequences were obtained from the GISAID database (https://www.epicov.org/epi3/frontend#5c1e82) or from internal CVL sequencing of clinical samples. Global analysis was performed using the NextStrain analysis tools (https://nextstrain.org/ncov/gisaid/global). The qPCR reactions were designed using the Genenious software package (www.geneious.com).

## 3. Results

### 3.1. Phylogenetic analysis of Omicron-specific mutations

The deletion in the spike gene at positions 22194-22196 and the deletion in the N gene at positions 28,362-28,371 were identified by Nextstrain analysis (https://nextstrain.org/ncov/gisaid/global) as unique for Omicron (**Figure 1**). Multiple alignment of available Omicron sequences obtained from GISAID (www.epicov.org/epi3/frontend#5c1e82) confirmed that the presence of these deletions, as well as the spike gene insertion in positions 221207-221215, were present in all sequenced Omicron samples. We therefore based our assays on these unique mutations.

**Figure 1.**
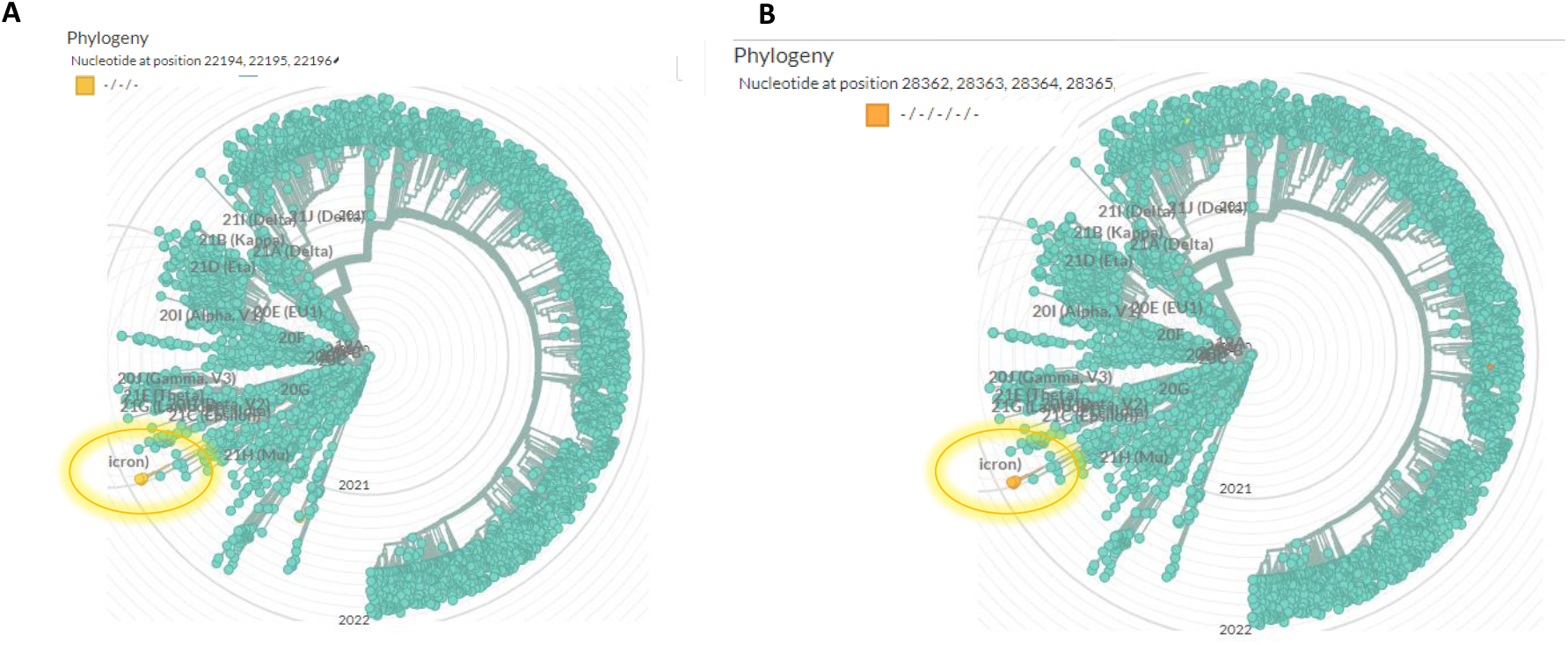
Global phylogenetic analysis showing the uniqueness of the spike 22194-22196 deletion (A) and the nucleocapsid (N) 28362-28371 deletion (B). the analysis was performed using Nextstrain (https://nextstrain.org/ncov/gisaid/global), based on sequences from the GISAID database (www.epicov.org/epi3/frontend#5c1e82). The lineage in which these mutations occur is circulated with a highlighted circle.

### 3.2. Development of negative N_del_ reaction

The assay targets the deletion at position 28362 of reference sequence NC_045512. The selectivity of the assay is accomplished by using the N1 R1a revers primer. Its annealing is strongly inhibited (>104-fold inhibition) in the presence of the deletion, thereby resulting in either absence of signal, or a 12-15 Cq values difference between the inclusive E-sarbeco reaction and the negative N_del_ reaction. The advantage of this assay is the use of widely available primer and probe developed by the CDC (3). **Figure 2** shows the amplification curves generated by testing A19 (“WT”), B.1.1.7 (Alpha), B.1.617 (Delta) and the suspected Omicron samples using the duplex assay containing the inclusive E-sarbeco reaction and the specific negative N_del_ reaction. The Cq values of each sample are shown in **Table 1**.

**Figure 2.**
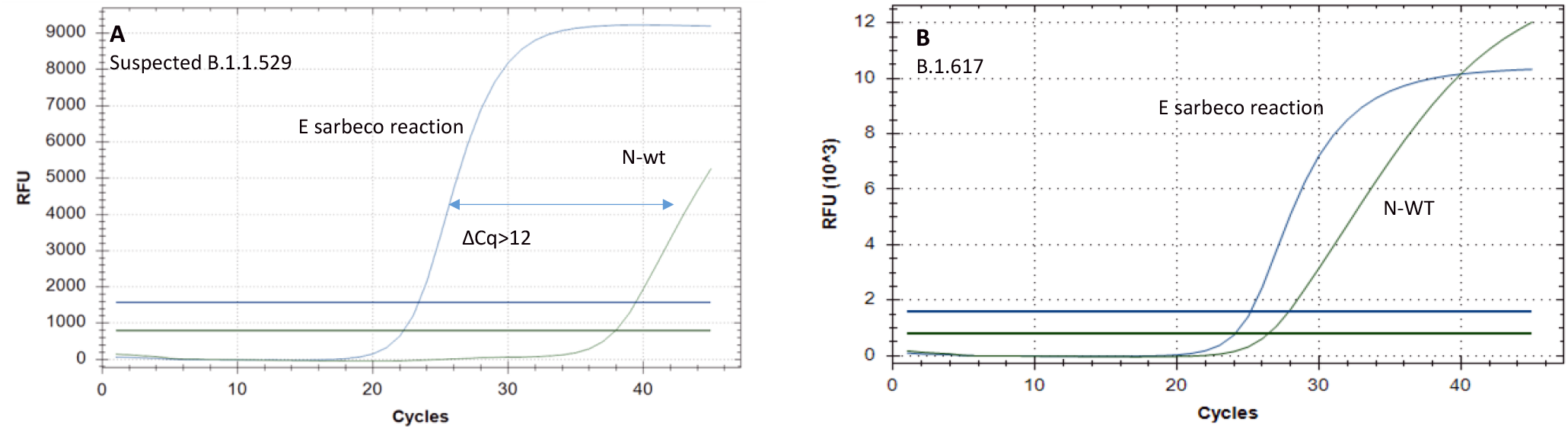
Amplification curves generated with the negative N_del_ reaction. (A) curves of suspected Omicron sample. (B) curves of a Delta sample.

**Table 1.** Cq values of each reaction for the different samples. The Cq difference between the Ndel reaction and the CoV19 reaction are shown in the “ΔCq” column.

| Sample | Cov19 E | $N_{del}$ | $\Delta Cq$ |
| --- | --- | --- | --- |
| WT | 30.43 | 31.44 | 1.01 |
| Alpha | 26.45 | 27.87 | 1.42 |
| Delta | 25.16 | 26.42 | 1.26 |
| Omicron | 23.38 | 37.96 | 14.58 |

### 3.3. Development of positive N_del_ reaction

In this reaction, the probe is annealing to the 9bp deletion at positions 28362-28371 of reference sequence NC_045512. Contrary to the negative N_del_ reaction, here the fluorescent signal is generated only in the presence of the deletion. **Figure 3** shows the amplification curves of serial dilutions of the first suspected Omicron sample obtained in Israel, and of A19, Alpha and Delta variant samples. **Table 2** details the Cq values obtained in this reaction.

**Figure 3.**
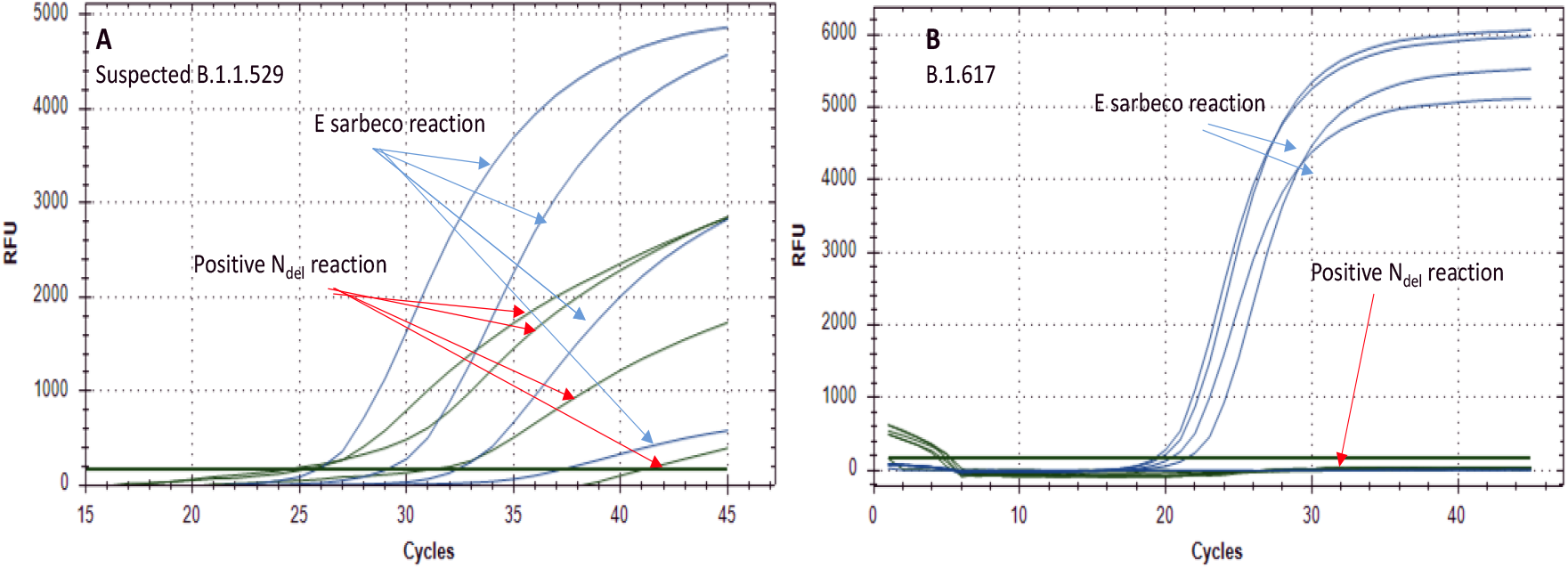
Amplification curves generated with the positive N_del_ reaction. (A) Serial dilutions of a suspected Omicron sample. (B) Clinical Delta samples. The curves of each reaction are indicated with corresponding arrows.

**Table 2.** Cq values obtained using the Positive N_del_ duplex assay.

| Sample | Cov19 E | Positive N<sub>del</sub> |
| --- | --- | --- |
| Susp. Omicron 10E-3 | 25.88 | 25.20 |
| Susp. Omicron 10E-4 | 29.12 | 28.45 |
| Susp. Omicron 10E-5 | 32.37 | 31.02 |
| Susp. Omicron 10E-6 | 37.33 | 41.03 |
| Delta 1 | 20.37 |  |
| Delta 2 | 19.62 |  |
| Delta 3 | 19.28 |  |
| Delta 4 | 21.47 |  |
| H<sub>2</sub>O |  |  |
| Neg RNA |  |  |

### 3.4. Development of the S_22194 indel_ reaction

This reaction targets the Omicron-unique deletion at positions 22194-22196 and insertion at positions 22207-22216 of reference sequence NC_045512. The probe in this reaction is complementary to the Omicron sequence and cannot bind to non-Omicron RNA. As described above, this assay was tested in a duplex with the SC-2 inclusive E-sarbeco reaction, testing the suspected Omicron sample and Delta samples. Figure 4 shows the resulting amplification curves and the Cq values are detailed in Table 3. It should be noted that these are preliminary results and the reaction should be further optimized for sensitivity and robustness. Its selectivity is, however, very specific and it allows clear identification of Omicron samples.

**Figure 4.**
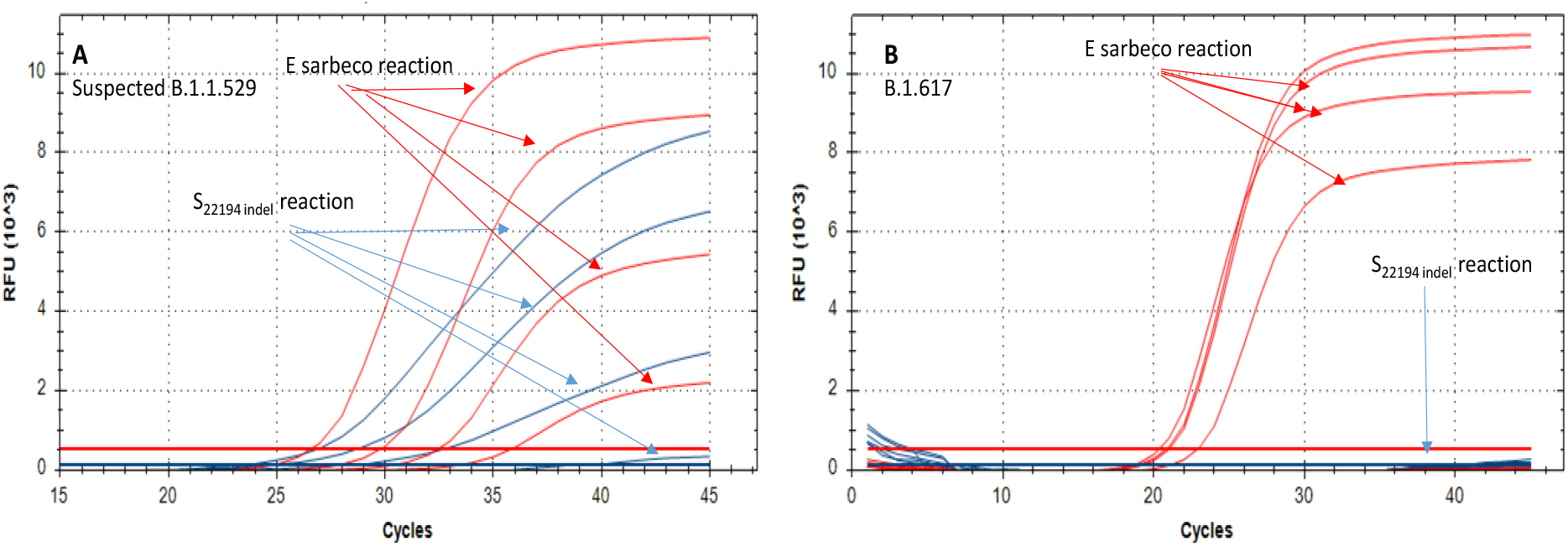
Amplification curves of the S_22194 indel_ reaction. (A) curves of dilutions of the suspected Omicron sample. (B) curves of four Delta samples. The curves of each reaction are indicated with corresponding arrows.

**Table 3.** Cq values obtained with the S_22194 indel_ reaction.

| Sample | $S_{22194}$ indel | Cov19 E |
| --- | --- | --- |
| Susp. B.1.1.529 10E-3 | 25.22 | 26.84 |
| Susp. B.1.1.529 10E-4 | 27.19 | 30.07 |
| Susp. B.1.1.529 10E-5 | 30.73 | 32.72 |
| Susp. B.1.1.529 10E-6 | 41.89 | 36.18 |
| Delta 1 |  | 21.21 |
| Delta 2 |  | 21.07 |
| Delta 3 |  | 20.57 |
| Delta 4 | 44.94 | 23.13 |
| H<sub>2</sub>O |  |  |
| Neg RNA |  |  |

### 3.5 Development of the NSP_del_ reaction

In this reaction, the forward primer is corresponding to the 9bp deletion at positions 11283-11292 of reference sequence NC_045512. When testing a deletion-positive sample, the Cq values obtained are similar to those obtained with the E-sarbeco reaction. In the absence of the deletion, the reaction is either completely, or strongly (>10^4^-fold) inhibited. Importantly, this deletion was detected in the Alpha, B.1.351 (Beta) and P.1 (Gamma) variants. However, since currently the only circulating variant in Israel and worldwide is Delta, this assay is sufficiently specific for identifying Omicron-suspected samples in Israel. Examination of complete genome sequencing in Israel from the last 3 months shows ∼0.03% incidence of this deletion in >3,000 examined samples (CVL unpublished data). Therefore, we assume that currently, this assay can be reliable in detecting Omicron-suspected samples. **Figure 5** shows the amplification curves of an Omicron-suspected sample, together with Alpha and Delta samples, tested with the E-sarbeco + NSP6_del_ reactions duplex. As expected, both the Alpha and Omicron-suspected samples were positive for the presence of the NSP6 deletion. The Delta samples were either negative for the NSP6_del_ reaction or weakly positive, with Cq difference of 12-14 cycles between the E reaction and the NSP6_del_ reaction. The Cq values of each reaction are detailed in **Table 4**.

**Figure 5.**
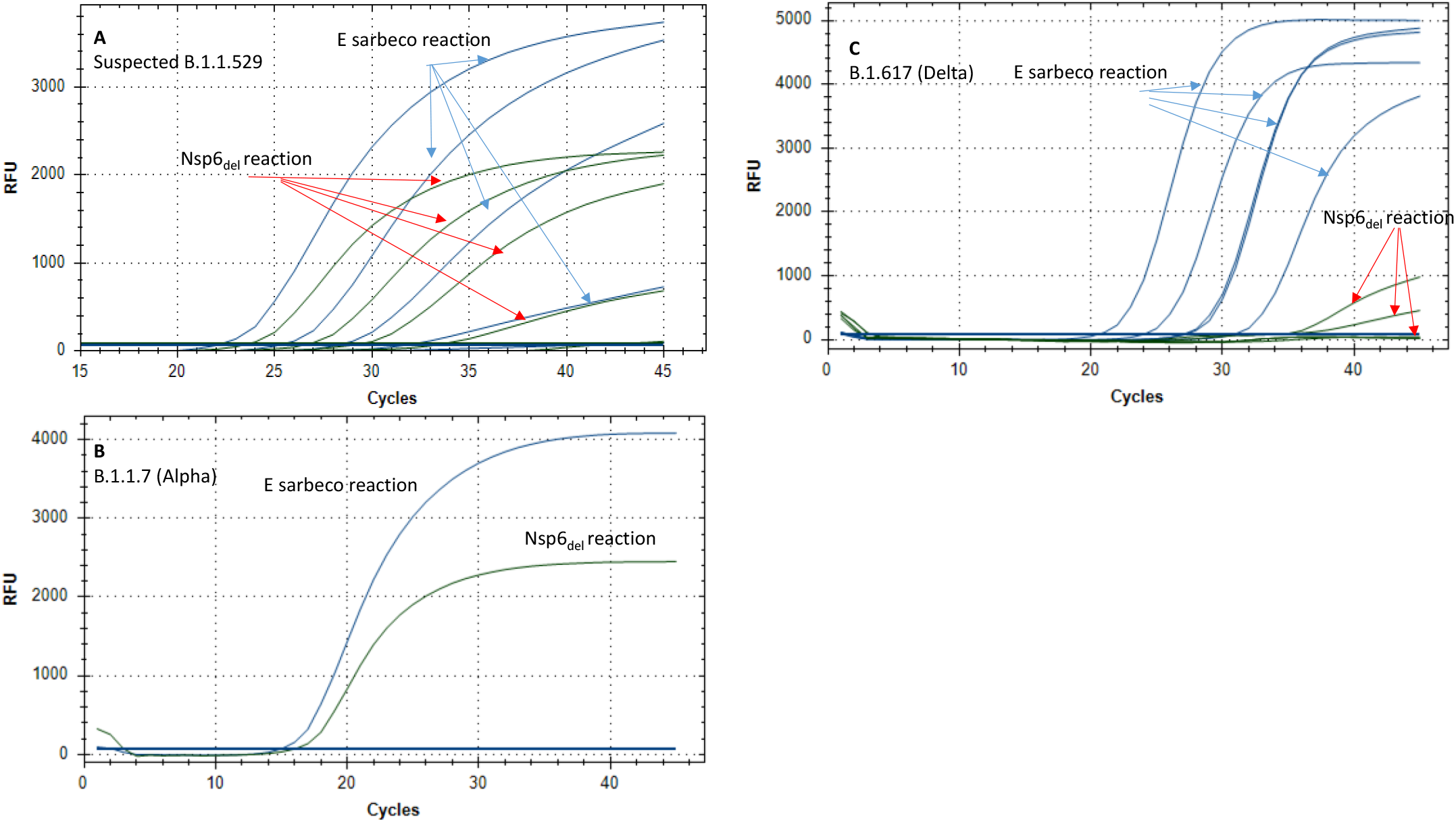
Amplification curves of the NSP6_del_ reaction. (A) Curves of serial dilutions of a suspected Omicron sample. (B) amplification of an Alpha sample. (C) Curves of five Delta samples. The curves of each reaction are indicated with corresponding arrows.

**Table 4.** Cq values obtained for the examined samples in the E-sarbeco + NSP6_del_ duplex assay. The Cq difference between the NSP6_del_ reaction and the CoV19 reaction are shown in the “ΔCq” column.

| Sample | COV19 E | nsp6<sub>del</sub> | $\Delta Cq$ |
| --- | --- | --- | --- |
| Susp. Omicron 10E-2 | 22.11 | 23.52 | 1.41 |
| Susp. Omicron 10E-3 | 25.28 | 26.63 | 1.35 |
| Susp. Omicron 10E-4 | 28.29 | 29.28 | 0.99 |
| Susp. Omicron 10E-5 | 31.92 | 33.48 | 1.56 |
| Susp. Omicron 10E-6 | 41.14 | 42.32 | 1.18 |
| Alpha | 15.02 | 16.14 | 1.12 |
| Delta 1 | 23.89 | 36.45 | 12.56 |
| Delta 2 | 26.9 |  |  |
| Delta 3 | 30.67 |  |  |
| Delta 5 | 20.4 | 34.12 | 13.72 |
| Delta 6 | 27.06 |  |  |
| CO-NEGATIVE |  |  |  |

## 4. Discussion

The four assays described herein are designed to help diagnostic laboratories to rapidly identify samples as Omicron. These protocols are fast, readily amendable for up scaling, and are far more simple and cost-effective compared with sequencing. The uniqueness of the mutations that the assays target render them highly accurate in identifying the suspected samples as Omicron, compared with other, less specific assays that detect more common mutations, such as the spike gene 69-70 deletion or the N501Y mutation. Indeed, the spike gene 69-70 deletion was found to exist in ∼1% of imported or circulating variants in Israel in the past 3 months (CVL unpublished data). The development of these assays was performed with a very limited number of positive samples, obtained from the first Omicron-suspected cases in Israel, which were verified as Omicron via sequencing. It is important to note that these results are preliminary and warrant further optimization. Due to the urgent need to make such discriminatory assays available to the international community, we made an effort to publish them as quickly as possible. It should be noted that the GISAID comprehensive database was important for the completion of this task in such a short time. The assays should be sufficiently robust to be implemented with reaction mixes from different manufacturers, such as Takara, PCRBIO, Thermo-Fisher Taqpath, and others. Reactions adjustments should be made, however, in the thermal profile, according to the specifications of each such product.

We plan to conduct a far more thorough and comprehensive validation for these assays, add endogenous control reaction, and publish our results in a proceeding report. We also welcome and comments and suggestions for improvements, should any such be available, from fellow laboratories.

## Data Availability

All data produced in the present study are available upon reasonable request to the authors

https://nextstrain.org/sars-cov-2/

https://www.gisaid.org/

https://www.cdc.gov/coronavirus/2019-ncov/lab/

## Ethical Statement

This study was conducted according to the guidelines of the Declaration of Helsinki, and approved by the Institutional Review Board of the Sheba Medical Center institutional review board (7045-20-SMC). Patient consent was waived as the study used remains of clinical samples and the analysis used anonymous clinical data.

